# The Impact of Host-Based Early Warning on Disease Outbreaks

**DOI:** 10.1101/2020.03.06.20029793

**Authors:** Mark A. Hernandez, Lauren E. Milechin, Shakti K. Davis, Richard A. DeLaura, Kajal T. Claypool, Albert J. Swiston

## Abstract

**Objective:** The detection of communicable pathogens responsible for major outbreaks relies on health care professionals’ recognition of symptoms manifesting in infectious individuals. Early warning of such communicable diseases before the onset of symptoms could improve both patient care and public health responses. However, the potential impact of such a host-based early warning system on containing the spread of an outbreak and in steering public health response is unknown.

**Methods:** We extend the deterministic SEIR (Susceptible, Exposed, Infectious, Recovered) model to simulate disease outbreak scenarios and to quantify the potential impact of a host-based early warning capability to mitigate pathogen transmission during an outbreak. In particular, we compare and contrast the performance of five different policies: Self-monitoring and reporting (baseline SEIR model), Quarantining the entire population, Quarantine-on-alert (with high sensitivity early warning), Quarantine-on-alert (with high specificity early warning), and Quarantine-on-alert (ideal early warning). We further evaluate these five policy options against four different outbreak scenarios with high or low disease transmission and high or low initial population exposures.

**Results:** For all scenarios, a quarantine-on-alert policy coupled with the near-ideal early warning capability reduces quarantine needs with only a small increase in the number of additional infections. The cost of a highly specific early detection system (i.e., a reduction in false alarms and thus quarantine costs) is an increase in additional infections relative to the near-ideal system. Conversely, a highly sensitive early detection system increases the percentage of the population in quarantine compared to both the ideal and high-specificity early detection system while also reducing the number of additional infections to nearly the numbers seen by quarantining the entire population a priori.

**Conclusions:** Our simulations demonstrate the utility of host-based early warning systems in controlling an outbreak under various outbreak conditions. Our tools also provide a simulation capability for evaluating public health policies enabling quantitative evaluation of their impacts prior to implementation.

## Introduction

The detection of communicable pathogens responsible for major outbreaks often relies on health care professionals’ recognition of symptoms manifesting in infectious individuals. Early warning of such communicable diseases before the onset of symptoms can potentially improve both patient care and public health responses. An example of such an early warning system is the PRESAGED (Presymptomatic Agent Exposure Detection) system [1] which uses host-based physiological signals to detect an individual’s exposure to pathogens, such as viruses and bacteria, before overt symptoms emerge and infectiousness is peak. In animal models, the PRESAGED algorithm has been shown to provide two to three days of early warning before the onset of incipient symptoms (e.g., fever), independent of the particular pathogen, exposure route, pathogen dose, or animal species [1]. These results are consistent with findings of Speranza et al. that show presymptomatic upregulation in biomarkers potentially linked to pathogen exposure around the same time in non-human primates exposed to Ebola [2].

Numerous efforts have evaluated epidemiological models to characterize the disease transmission dynamics and the effectiveness of public health interventions of past outbreaks [9–13], including effort by Chowell et al [14] that examines the impact of a hypothetical early diagnostic capability (based on advancements in bioassay tests) for containing the spread of Ebola. We posit that a comprehensive simulation capability is essential for assessing the potential impact of a host-based early warning system.

We have developed a series of epidemiological models that quantify the potential impact of a host-based early warning capability to mitigate pathogen transmission during an outbreak. Our epidemiological models reflect a variety of traditional public health policies related to nonpharmaceutical interventions, such as quarantine and patient isolation, and novel policies enabled by host-based early warning capabilities, such as the PRESAGED system [1]. For each policy-dependent model, we simulated outbreak scenarios and calculated the size of the outbreak (total number of infections) and the operational burden (total number of lost duty days resulting from quarantine or isolation). These metrics were used to understand the trade space for the different policies. Our simulations demonstrate the utility of host-based early warning systems in controlling an outbreak under various outbreak conditions.

## Models for Disease Outbreak Simulation

### The SEIR Model

One of the most common epidemiological models for simulating disease outbreak scenarios is the deterministic SEIR model. This approach splits a given population into separate compartments defined by their relationship to a disease outbreak [15]:

1. Susceptible: healthy individuals who can be exposed to the pathogen
2. Exposed: individuals who are in the incubation phase; they have been exposed to the pathogen but are not yet showing symptoms and are not infectious
3. Infectious: individuals who are infectious to the susceptible population and will eventually display overt symptoms
4. Recovered: those who have recovered from illness and acquired immunity to further infection

These compartments are then linked with a system of ordinary differential equations (ODEs) to characterize how individuals transition into and out of each compartment over time. A variety of scenarios can be simulated by changing the rate parameters of the ODEs linking the population compartments. Furthermore, this approach allows quantitative projections of how many people are exposed to a pathogen and become sick under different outbreak conditions.

While the SEIR model is mathematically rigorous and often has good predictive utility during an outbreak, the SEIR model requires several assumptions. First, the model assumes there is a fixed population *N*, with no births or deaths other than those resulting from the infectious disease. Second, it assumes the population is homogeneously mixing, that is the transmission between any two individuals is equally likely. Third, the model assumes that exposed individuals become infectious after a fixed incubation period, thus not accounting for individual variability of disease progression for the young, elderly, or immunocompromised. Finally, the model assumes that all recovered individuals are immune to further infection and thus do not re-enter the susceptible class. While any of these assumptions may fail to hold in particular contexts, abiding by them allows for greater mathematical tractability and offers similar relative output trends.

### Baseline SEIR Model

The SEIR model consists of a system of four ODEs that describe the rate of change of individuals in each compartment over time (Figure 1). The differential equations for Susceptible, *S*_*t*_, Exposed, *E*_*t*_, Infectious, *I*_*t*_, and Recovered, *R*_*t*_ are as follows, where dependence on time, *t*, is omitted in the notation below for simplicity:

**FIGURE 1.**
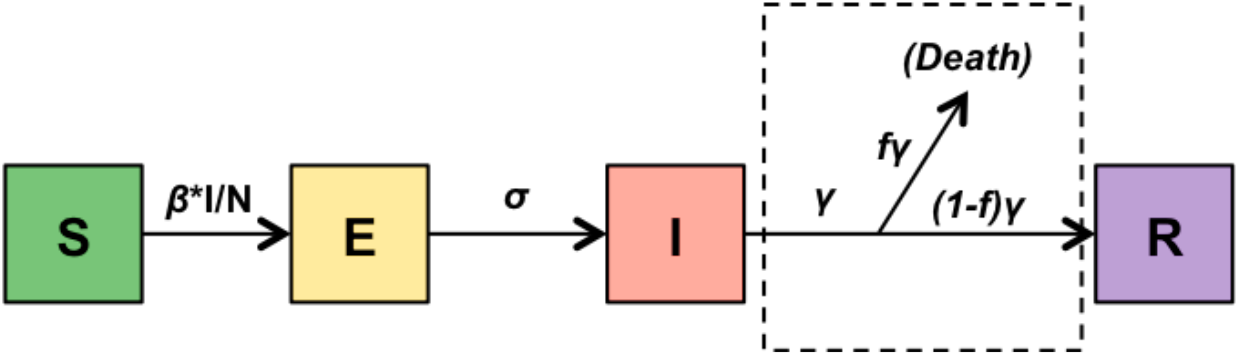
In this schematic diagram of the SEIR model, each box represents a compartment of the population, and arrows show the progression of individuals through those compartments. Expressions above arrows are rate coefficients showing progression through compartments. Note that the dashed boxed portion, indicating individuals leaving the population because of death, will not be included in future diagrams for simplicity.

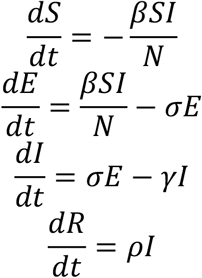

Table 1 describes all of the parameters we use for our models. At the start of the model, a subset of the population is exposed to a pathogen (*E*_*0*_), and the remainder are susceptible to infection. Susceptible individuals enter the exposed compartment at a rate of *^βSI^/_N_*, which is known as the normalized transmission rate. The parameter *β* is the contact rate, which accounts for how often susceptible-infectious contacts result in a susceptible individual becoming exposed to the pathogen. Exposed individuals become infectious at a rate of σ, which is the inverse of the incubation period. Infectious individuals stay infectious at a rate of *γ*(the inverse of the infectious period) until they recover or die. We define the recovery rate, *ρ* as (1-f) *γ* where the case fatality rate *f* is the proportion of infected individuals who die from the disease. Note that we do not specify a mortality compartment in this analysis, though for some pathogens spread through the handling of remains (such as Ebola during the 2014 West Africa outbreak), this compartment would be a critical addition.

**TABLE 1.**
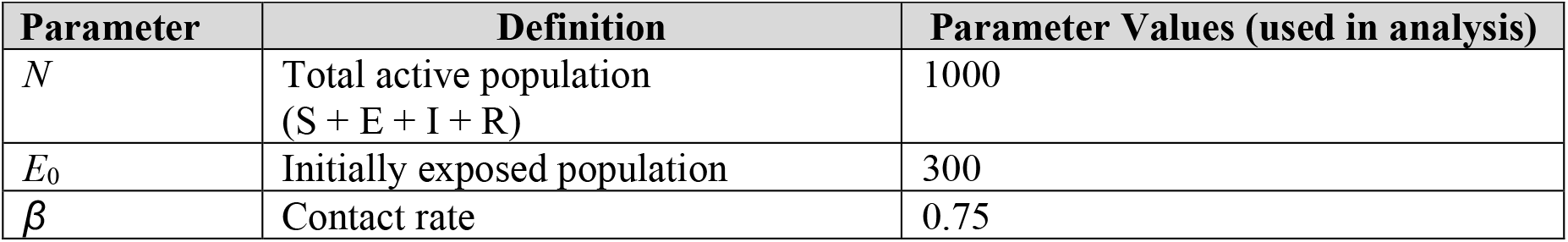
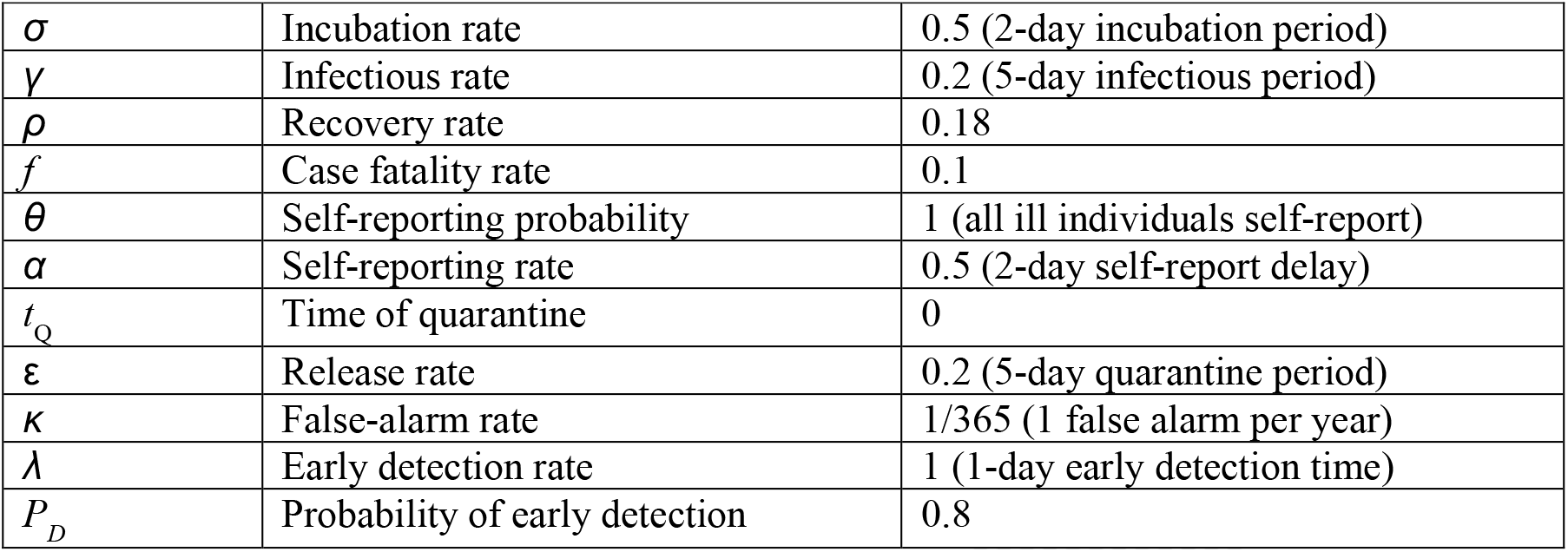
Parameter Definitions and Values for Baseline and Policy-Dependent Models.

Another important component of the SEIR model is the basic reproduction number *ℛ*_0_which is often included as a basic property of any given pathogen [15]. *ℛ*_0_ represents the average number of additional infections caused by each infectious individual, assuming there are no control interventions. In a fixed population represented in the SEIR model, the reproduction number can be calculated from parameters in the model as *ℛ*_0_= *^β^/_γ_*.

The solutions of the SEIR model are functions of the number of individuals in each compartment with respect to time, i.e., *S*_*t*_, *E*_*t*_, *I*_*t*_, and *R*_*t*_. We solve this numerically by using an ODE solver in MATLAB®.

An example output of the full SEIR model is shown in Figure 2a. Here, we begin with a population of *N* = 1000 people (roughly equivalent to a large battalion), and consider a scenario where 300 individuals are exposed to some pathogen at time, *t* = 0(*S*_*t*=0_ = 700,*E*_*t*=0_ = 300,*I*_*t*=0_ = *R*_*t*=0_ =0). If we assume a particular infectious pathogen has a contact rate, *β* =0.75, with an incubation period of two days, σ=0.5, and an infectious period of five days, *γ*=0.2(similar properties to a highly contagious flu virus), the solution to the SEIR model shows that nearly the entire population will contract the disease over the course of a month. Figure 2b focuses on the number of cumulative infections, a metric for assessing the overall size of the outbreak over time. This baseline condition, in which no public health policy is in effect, represents the most severe outcome from an infectious disease outbreak.

**FIGURE 2.**
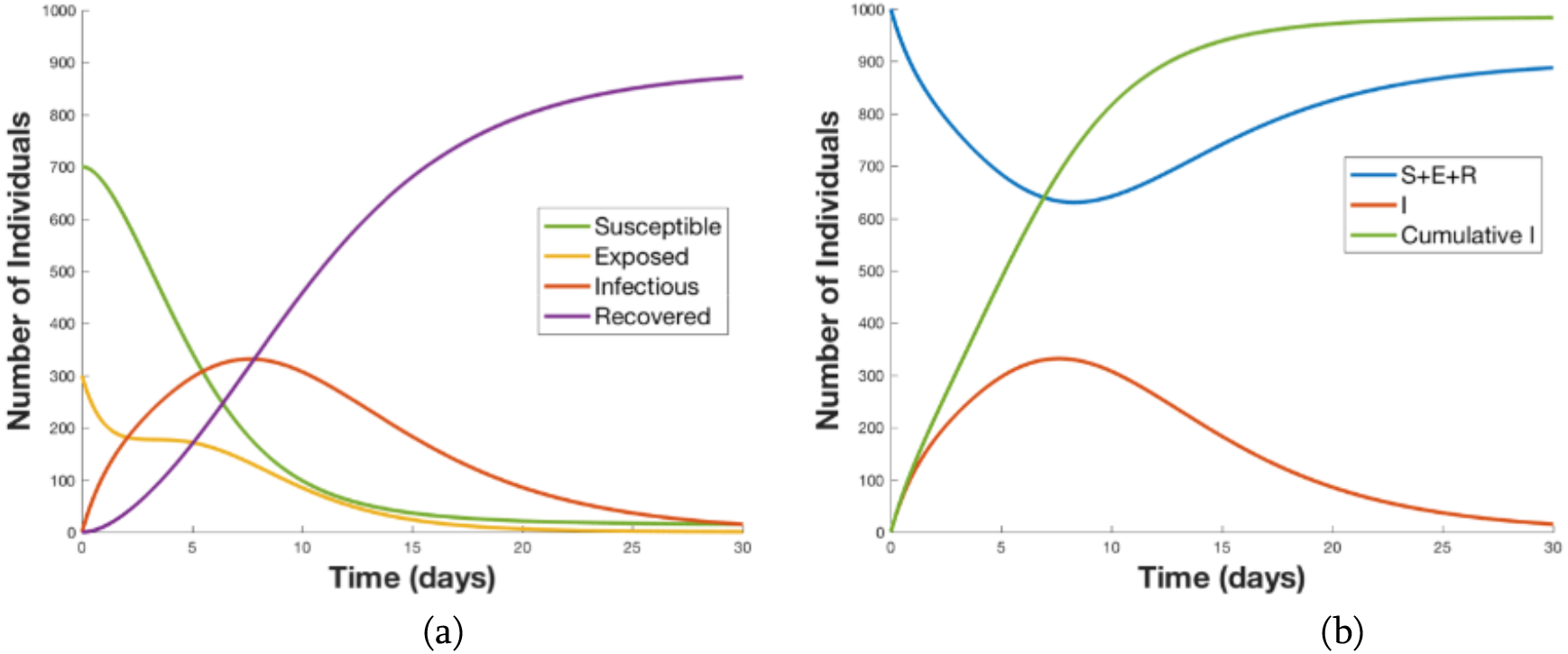
A numeric solution for the baseline SEIR model shows the population of each compartment versus time (a), which indicates that for an initial exposure of 300 individuals, nearly the entire population will eventually fall ill; the cumulative number of infections (Cumulative I) versus time is shown in (b).

### Policy-Dependent SEIR Models

In the previous section, we considered the standard SEIR model, which describes disease spread *without* any public health interventions. To quantitatively evaluate an outbreak scenario with additional measures, the parameters and compartments of the SEIR model can be modified to reflect policy choices or new early warning technologies. Two policies currently considered as standards in handling possible outbreaks rely on self-monitoring to implement voluntary isolation or quarantining all individuals that may have been exposed to a pathogen. Isolation applies to individuals who are already ill, whereas quarantine applies to individuals who may have been exposed but have not yet shown symptoms. The first policy, self-monitoring and reporting, which assumes individuals self-report when they develop symptoms, allows for additional infections during the time delay between the onset of symptoms and isolation; however, it does not account for the logistical and financial cost of quarantine. The second policy, quarantining the entire population, is effective in reducing additional transmissions but has a prohibitively onerous costs for large populations. In an effort to explore the impact of a host-based early warning system, we modified the SEIR model to simulate a third policy, quarantine-on-alert, in which individuals are only quarantined when prompted by a host-based early warning system. We hypothesized that the early notification of incipient illness will allow for reductions in pathogen transmission while minimizing the number of quarantined individuals. We then assessed each of these three new policy-dependent SEIR models.

### Self-Monitoring and Reporting: Isolation after Symptoms

In the course of an infectious disease outbreak, most individuals tend to self-monitor for symptoms of the pathogen. If individuals start to develop symptoms, they can self-report to a medical facility and may be immediately isolated until they recover. A new compartment, isoLated (*L*), was added to the SEIR model to reflect this symptomatic and infectious population, which has limited contact with the rest of the population and therefore reduced transmission potential (Figure 3). The self-reporting probability *θ* is defined as the proportion of symptomatic individuals who are compliant in reporting their symptoms and enter the *L* compartment. The self-reporting rate α is derived from the delay between developing symptoms and self-reporting (i.e., the inverse of the average time delay). For our model, we assumed that isolation is 100 percent effective in preventing transmission, and individuals in the *L* compartment are unable to infect the susceptible population. While a self-monitoring policy reduces transmission rates by isolating the self-reported sick, both the latency α and an imperfect self-reporting probability (*θ*<1) can lead to opportunities for transmission.

**FIGURE 3.**
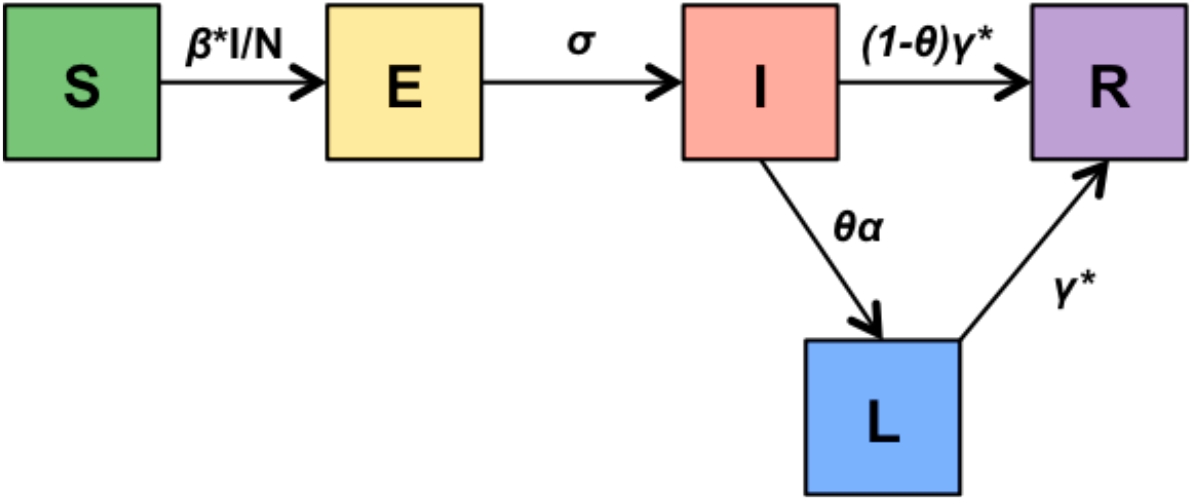
The model diagram for a self-monitoring and self-reporting policy includes an additional compartment (*L*) for individuals who are isolated after developing symptoms. Note that fatalities are left out for simplicity.

Outputs for the SEIR model with self-reporting and isolation are shown in Figure 4. This policy addition leads to both fewer total individuals contracting the disease and fewer individuals acutely symptomatic at the height of the outbreak. The new isolated compartment *L*, however, has other costs associated with lost duty days, mandated isolation, medical facilities, and treatment. The choices of self-reporting probability and rate are critical in this scenario, and, in reality, may have such broad distributions that relying on this policy in an acute outbreak may do little to prevent additional infections.

**FIGURE 4.**
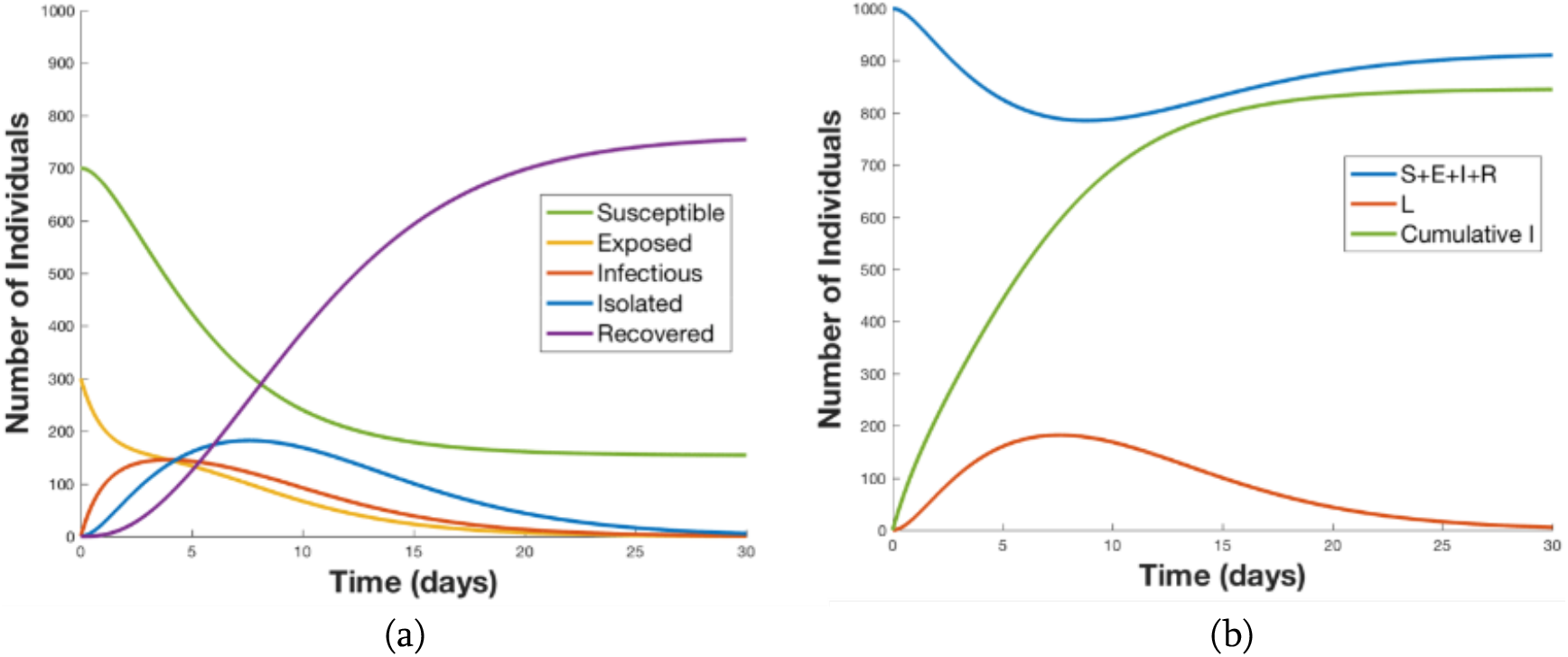
In this numeric solution for our SEIR model with a self-monitoring and isolation policy enacted, the population of each compartment versus time is shown in (a), and the active working (S + E + I + R) and inactive (L) populations versus time with the cumulative infections versus time are shown in (b). The self-monitoring policy has effectively reduced the total number of disease cases and blunted the outbreak’s peak severity (number of cases in I and L peak at approximately 7 days).

**FIGURE 5.**
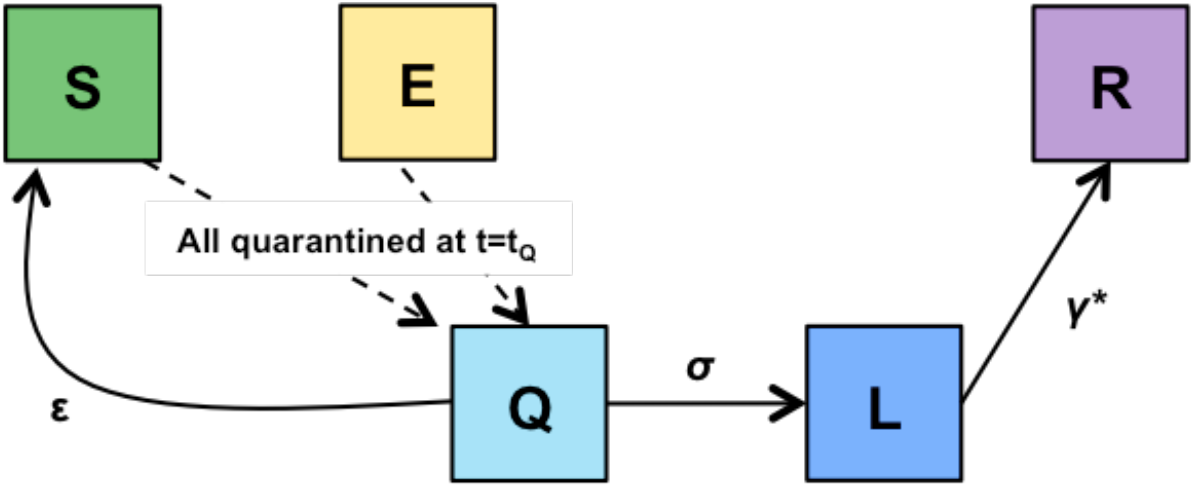
The model diagram for a quarantine-all policy moves all susceptible and exposed individuals to the quarantined compartment (Q) at the time t0, Quarantined individuals who do not develop symptoms are returned to the susceptible compartment after a maximum incubation time.

### Quarantining the Entire Population

Another possible public health response to an outbreak is to quarantine currently healthy individuals with some likelihood of pathogen exposure. In outbreaks where a large population subset has some exposure likelihood, a quarantine-all policy would separate all individuals in the population from contact with one another. Our model assumes that quarantined individuals, represented by the *Q* compartment, are monitored and immediately isolated once they develop symptoms (Figure 5). If quarantined individuals do not become symptomatic after the maximum incubation time of the suspected pathogen (often several weeks), they are released and re-enter the *S* compartment at a rate of ε (the inverse of the maximum incubation time). As shown in Figure 6, the quarantine-all policy eliminates opportunities for further pathogen transmission; however, it also results in the quarantine of healthy individuals who have not been exposed, contributing to immense productivity losses, extreme logistic burdens associated with providing accommodations for an entire population, and acute civil rights concerns. Furthermore, the assumption that transmission is zero within the *Q* compartment may not be realistic because transmission for some pathogen may occur before overt symptoms and isolation. This quarantine-all policy could result in the illness of people who, if not for the quarantine, would never have been exposed to the pathogen.

**FIGURE 5.**
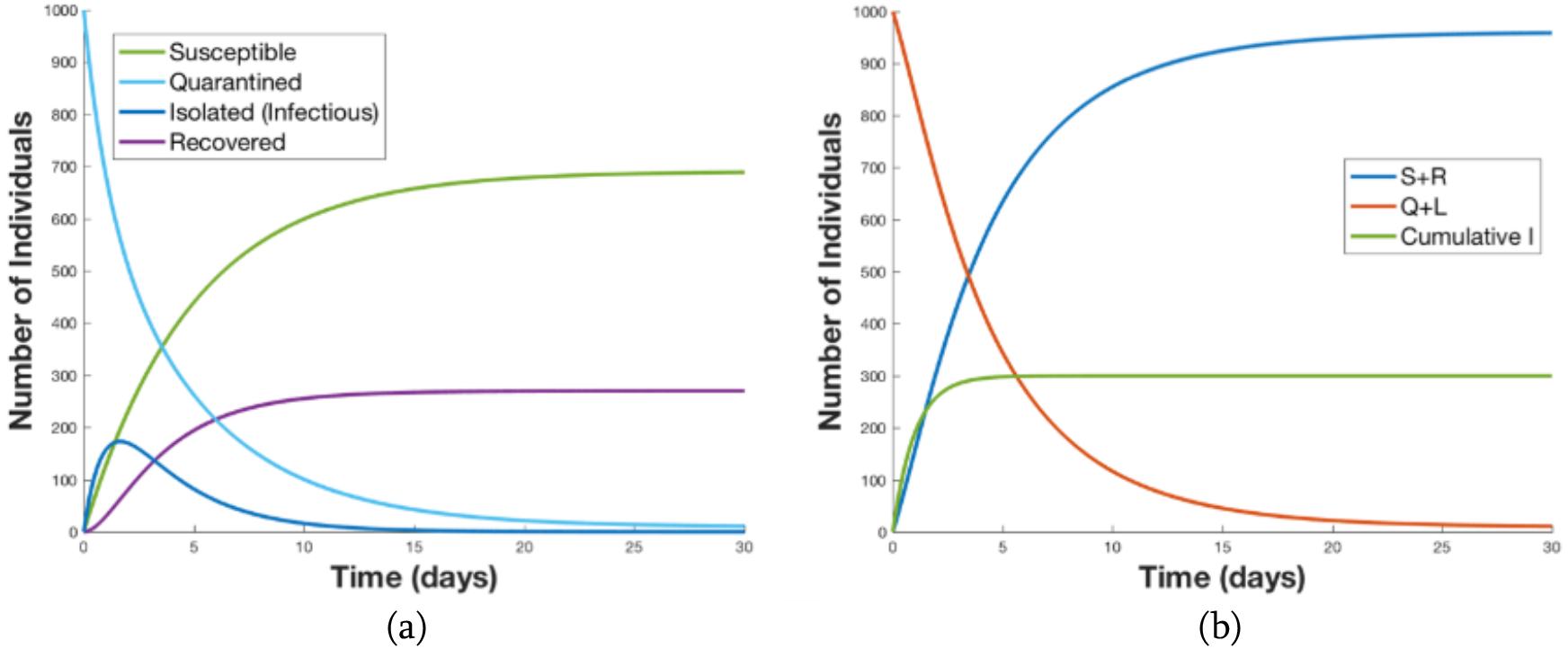
In the numeric solution for our SEIR Model with a quarantine-all policy enacted, the population of each compartment versus time, which notably does not include an exposed compartment, is shown in (a). In (a) are individuals suspected to be exposed and quarantined initially, until they either fall ill (and are isolated) or are released after the quarantine duration (21 days). The active working (S + R) and inactive (Q + L) populations versus time with the cumulative infections versus time are shown in (b). The quarantine-all policy has very effectively reduced the total number of disease cases but has vastly increased the burdens of quarantine and isolation.

**FIGURE 6.**
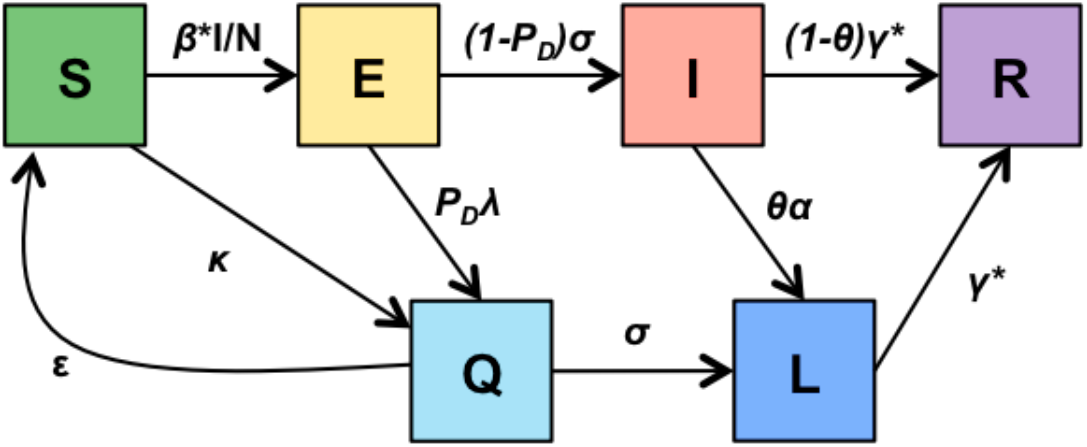
This hybrid model diagram illustrates the final quarantine-on-alert policy. The performance of the PRESAGED-like early warning system is now explicitly added, including the probabilities of early detection and false alarm, as well as an expected early detection time before symptom onset and infectiousness. This hybrid between quarantine-all and self-monitoring with isolation seeks to leverage the epidemic-limiting behavior of both policies while reducing the cost and burdens of each.

### Quarantine on Alert

The third model we explored simulates a novel policy that could potentially be enabled by future host-based early detection capabilities. Here, a host-based early warning system could prompt a quarantine only on alert when the system detects pre-symptomatic signs of exposure. We integrated the early warning performance metrics from our PRESAGED algorithm—population-wide probability of early detection, daily false-alarm rate, and early detection time—into a base SEIR model. As shown in Figure 7, a host-based early warning system would trigger exposed individuals to transition into quarantine at a rate characterized by the product of the parameters *λ* and *P*_*D*_. The parameter *λ* is an early detection rate defined as the inverse of the system’s average early detection time for a given pathogen. The parameter *P*_*D*_ is the population-wide probability of early detection, i.e., the fraction of the exposed population that will present early detections of pathogen exposure prior to becoming infectious. Missed detection cases, when the system fails to produce an early warning, occur with a probability of *1 – P*_*D*_ across the exposed population. For this policy, false alarms of the host-based early warning system would cause healthy individuals to be incorrectly quarantined. Our model takes this notion into account by adding a transition rate from the susceptible to the quarantined compartment, characterized by a daily false-alarm rate *κ*, the probability of an individual’s early warning system presenting a false alarm on a given day.

**FIGURE 7.**
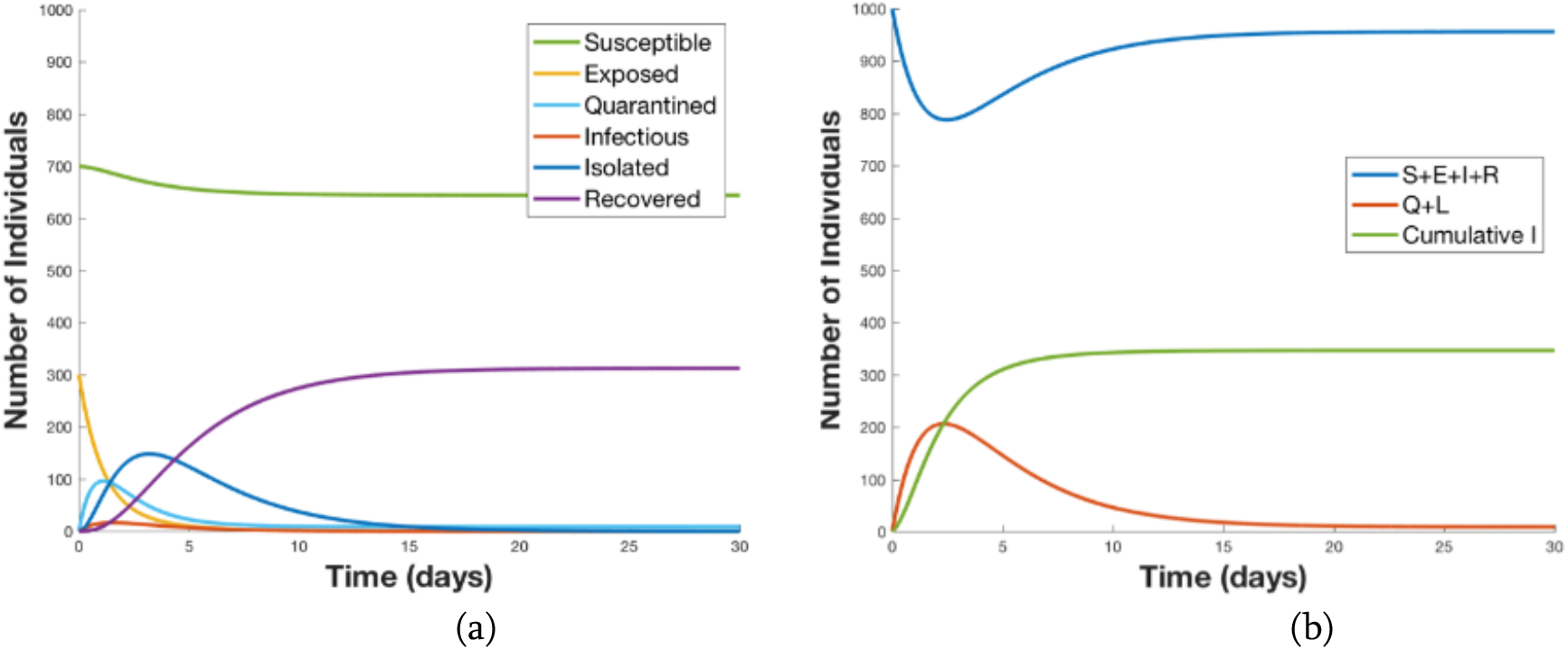
This numeric solution for our SEIR model with a quarantine-on-alert policy enacted shows the population of each compartment versus time (a); the plot shows how such an early warning technology could limit the outbreak size. As the initially exposed individuals receive alerts, they enter the quarantine-on-alert compartment and are unable to infect the susceptible compartment. The active working (S + E + I + R) and inactive (Q + L) populations versus time are shown with the cumulative infections versus time in (b). The quarantine-on-alert policy has effectively reduced the additional disease cases and has largely limited those in quarantine to those individuals who were initially exposed.

While the self-monitoring and quarantine-all policies present clear trade-offs between cumulative number of infections and number of people isolated or quarantined, this quarantine-on-alert policy offers a hybrid approach that could optimize for both. Quarantine-on-alert vastly reduces quarantine costs by selectively identifying likely exposed individuals through physiologically based predictions. Furthermore, once quarantined, these high-risk individuals can be immediately isolated, thus limiting the opportunities of additional infections and enabling more focused medical care.

### SEIR Model Reproduction Numbers by Policy Choice

Modifications made to the baseline SEIR model result in changes to the reproduction number (*ℛ*) associated with each policy-dependent SEIR model [16]. Our example scenario assumes that every symptomatic person will self-report and thus the self-reporting probability, *θ*, is 1. Table 2 summarizes equations to calculate *ℛ* values for each policy, the *ℛ* values given the parameter values in Table 1, and the cumulative cases of infection for each policy choice after 50 days. The policies with the lower *ℛ* values (quarantine-all and quarantine-on-alert) are associated with minimal pathogen spread, while the policies with higher *ℛ* values (self-monitoring and baseline) yield a greater number of infection cases. The number of cumulative infections as the ODE reaches equilibrium illustrates how the reproduction number would affect the final size of the outbreak. Note that the number of cumulative infections for baseline saturates around the population size (*N* = 1000) when the number of individuals in the susceptible compartment is exhausted.

**TABLE 2.** Reproduction Number (*ℛ*) Equations and Nominal ℛ Values for Our Example Scenario.

| Policy | $\mathcal{R}$ Equation | $\mathcal{R}$ Value | Cumulative Infections after 50 days |
| --- | --- | --- | --- |
| Baseline | $\mathcal{R}_0 = \frac{\beta}{\gamma}$ | 3.75 | 986 |
| Self-monitoring | $\mathcal{R}_0 = \frac{\beta}{\alpha}$ | 1.5 | 846 |
| Quarantine-all | $\mathcal{R} = 0$ | 0 | 300 |
| Quarantine-on-alert | $\mathcal{R} = \frac{\beta}{\alpha} \frac{(1 - P_D)\sigma}{(1 - P_D)\sigma + P_D\lambda}$ | 0.167 | 347 |
| In a population of $N = 1000$, the number of initially exposed individuals is $E_0 = 300$. The nominal $\mathcal{R}$ Value and Cumulative Infections were calculated using parameter values from Table 1. | | | |

#### Policy Trade-Space Analysis

The policy-dependent SEIR models provide a foundation for a trade-space analysis of different quarantine, isolation, and treatment (QIT) policies as a function of disease transmission characteristics, exposure scenarios, and performance of early warning systems. The analysis captures both the potential benefits (reduction in quarantine costs, more focused use of medical resources) and risks (increase in infections) for each policy.

To demonstrate the potential utility of the early warning–enabled quarantine-on-alert model in a more comprehensive QIT policy analysis, we compared outcomes over a range of disease transmission rates, initial exposure conditions, and early warning performance parameters. We varied these parameters in simulations of a mass exposure to an infectious pathogen occurring in a population of 1,000 people. All scenarios were simulated over 50 days. Because we fixed the exposure time for all cases to *t* = 0, rather than using the quarantine release rate ε, all individuals in quarantine were returned to the susceptible compartment at *t* = 21 days (the maximum incubation period).

We defined policy outcomes via two metrics derived from the policy-dependent SEIR model outputs as shown in Figure 9. The first, lost duty days, is the percentage of total number of days of work productivity that are lost because of quarantine and isolation in the 50-day simulation; this percentage is proportional to the integral of the curves in Figure 9a. The second metric, cumulative infections, is the percentage of the population that has been infected by the end of the simulation (Figure 9b). These two metrics characterize a trade space for evaluating QIT policies under different circumstances, as any measure that reduces the number of people in quarantine and isolation may be expected to increase the likelihood of infection in the population. While these metrics are helpful for evaluating the impact of various policy choices, they are by no means comprehensive and particularly do not consider the financial or other costs associated with quarantine.

**FIGURE 9.**
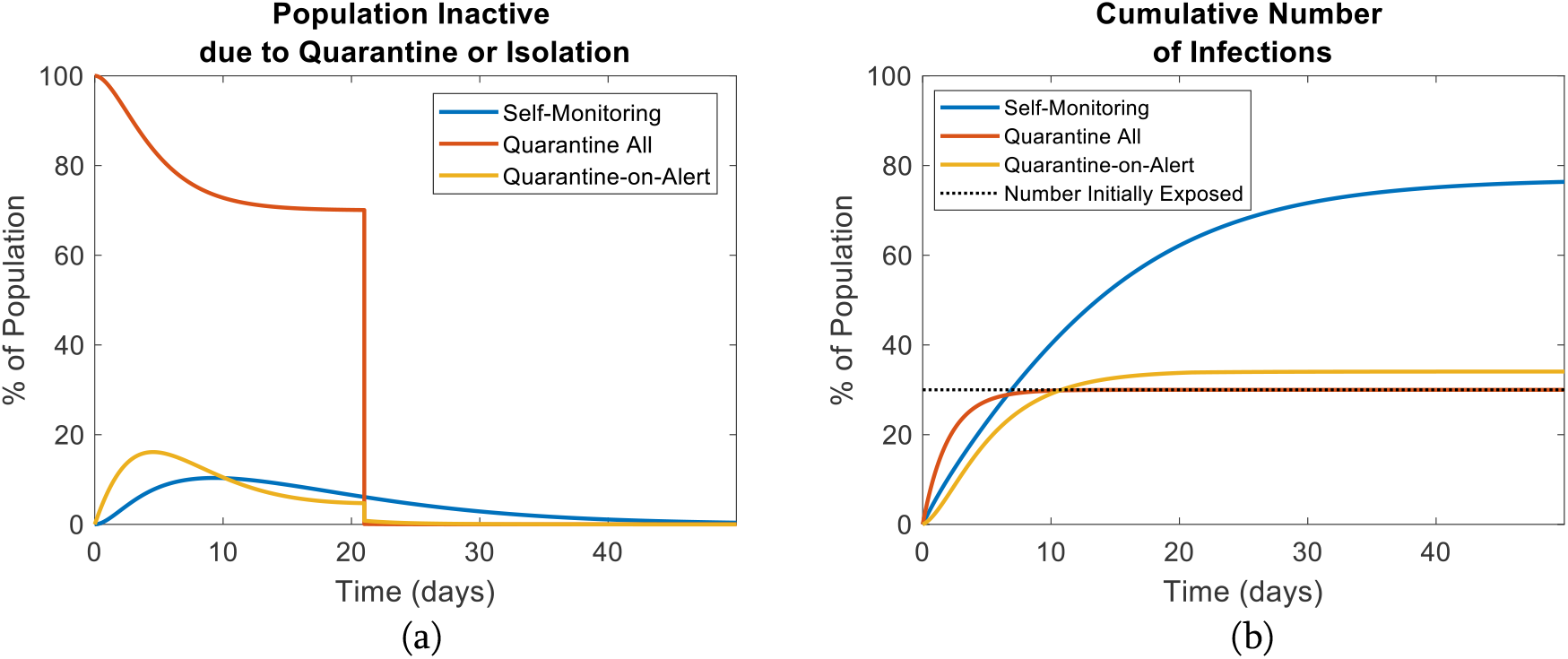
The model outputs compare consequences of self-monitoring, quarantine-all, and quarantine-on-alert outbreak response policies. In (a), the plot shows the number of inactive individuals that are unable to work due to quarantine and isolation (Q + L for quarantine-all and quarantine-on-alert, only L for self-monitoring because it does not utilize quarantine) versus time. The sharp drop-off for quarantine-all represents the release of all quarantined individuals after 21 days. In (b), the plot shows the cumulative infections versus time. Any infections above the dotted line (number of individuals initially exposed at *t* = 0) indicate additional transmission of the pathogen.

Using these metrics, we evaluated five QIT policy sets: quarantine-all, isolate upon self-reporting, and quarantine-on-alert with three different levels of early detection performance— high sensitivity (*P*_*D*_ = 0.8, κ =0.10), high specificity (*P*_*D*_ = 0.4, κ = 1/365), and a near-ideal early-detection system (*P*_*D*_ = 0.8, κ = 1/365). These five QIT policy options were then tested against four different outbreak scenarios with high or low disease transmission (where β = 0.6 or 0.3, respectively) and high or low initial population exposures (where *E*_*0* =_ 600 or 50, respectively). Figure 10 shows the QIT policy trade-offs for these 20 independent outbreak and policy combinations.

**FIGURE 10.**
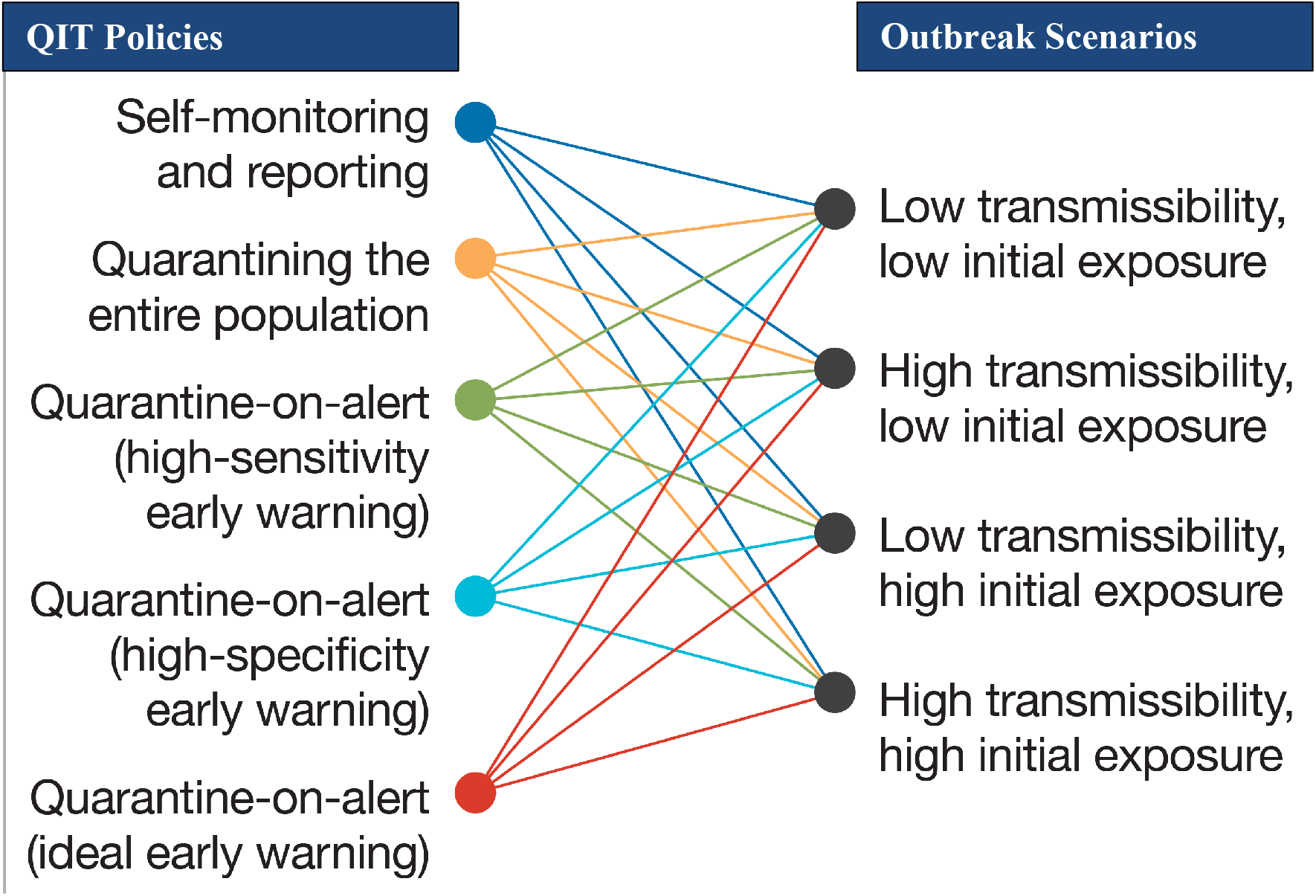
Simulations are run on each combination of QIT policy and outbreak scenario, resulting in 20 independent conditions for the trade space simulations.

We found that for all scenarios, a quarantine-on-alert policy coupled with the near-ideal early warning capability reduced quarantine needs with only a small increase in the number of additional infections. The cost of focusing on a high-specificity early detection system (i.e., a reduction in false alarms and thus quarantine costs) was an increase in additional infections relative to the near-ideal system. Conversely, a high-sensitivity system increased the percentage of the population in quarantine compared to both the ideal and high-specificity early detection system while reducing the number of additional infections to nearly the number seen by quarantining the entire population a priori.

For scenarios with low initial exposure, the impact of early warning–enabled QIT policies varied dramatically with transmissibility. For low-transmissibility and small initial exposures (Figure 11a), an early warning capability demonstrated the least utility as the outbreak was effectively contained with just self-monitoring. However, for a pathogen with high transmissibility (Figure 11b), the cumulative infections were reduced significantly with the notional early warning systems. A high-specificity system offered a particularly promising result, showing comparable infection reduction to the near-ideal system while also minimizing the number of days lost to quarantine.

**FIGURE 11.**
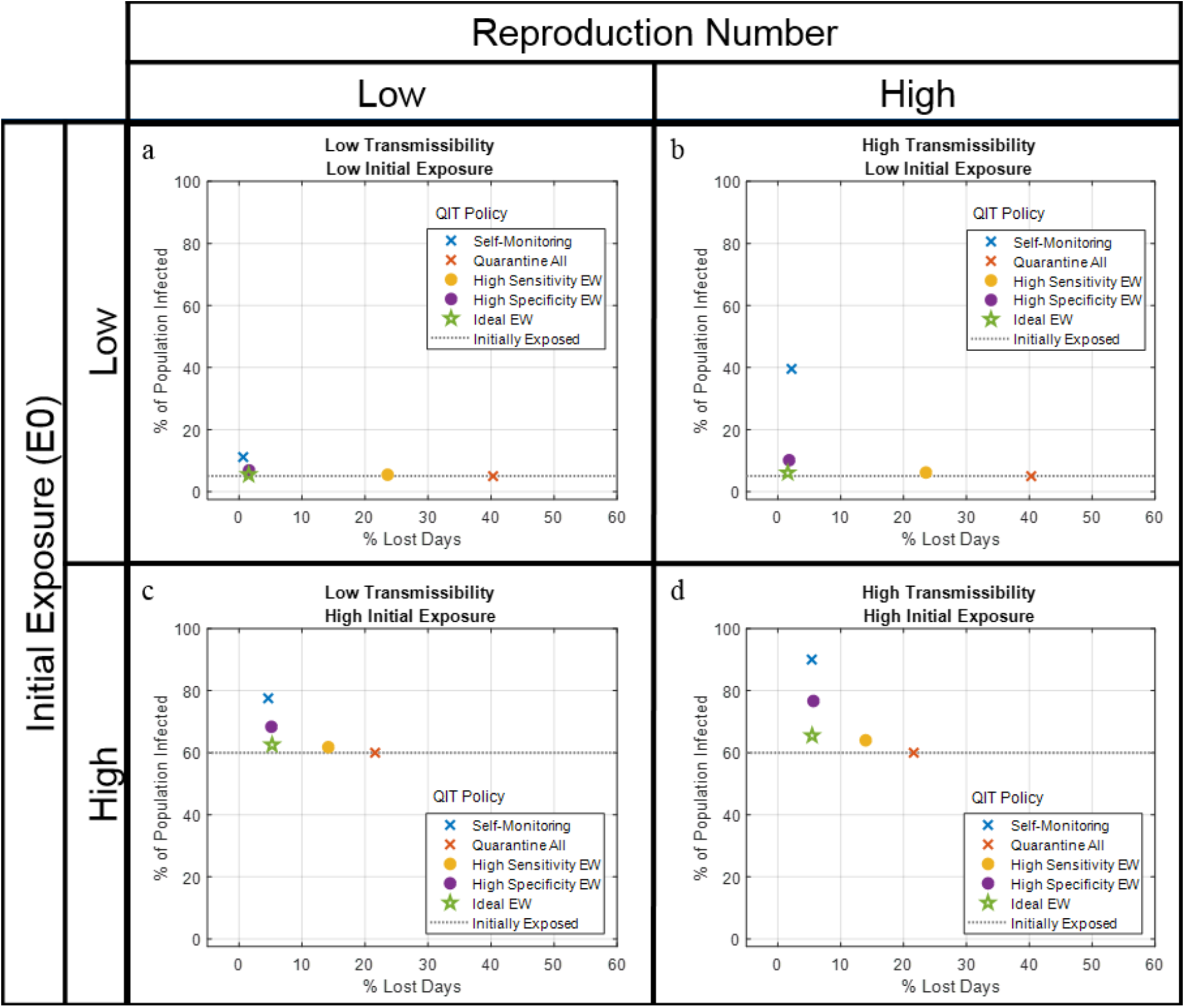
The trade space for the percentage of population infected versus percentage of lost duty days compares the augmented policies: self-monitoring, quarantine-all, and quarantine-on-alert with three levels of early warning performance (high sensitivity, high specificity, and ideal early warning). Four situations were considered: low-transmissibility pathogen with a low initial exposure (a), high-transmissibility pathogen with a low initial exposure (b), low-transmissibility pathogen with a high initial exposure (c), and high-transmissibility pathogen with a high initial exposure (d).

In the scenarios of high initial exposure (Figure 11c and Figure 11d), the costs of imperfect early detection were more pronounced compared to those for scenarios of low initial exposure. Because a high initial exposure greatly increased the likelihood of infectious individuals transmitting the pathogen to those susceptible, even a short delay in isolating infectious individuals lead to more infections. The reduction of cumulative infections for all notional early warning systems, relative to the self-monitoring policy, was less prominent than in scenarios of low initial exposure as the initial exposed population was close to saturation. In effect, for these extreme cases of mass exposures, the more aggressive policies (i.e., a high-sensitivity early detection system or quarantining everyone) may be more effective.

#### Discussion and Future Work

Our results on the early warning–enabled and policy-dependent SEIR models allows for a quantitative analysis of the QIT trade space and provides potential guidance on priorities for the future development of early warning technology. However, this illustration of QIT risk analysis captures only a subset of the factors that must be considered in the formulation of a rational, effective QIT policy. The context of the scenario will ultimately inform where the ideal operating point would be. For example, the number of total infections may be interpreted differently depending upon the virulence of the disease. The tolerance for new infections may be low if the consequences are high, such as if the infection is almost always fatal or is associated with severe symptoms and long-term complications. Additionally, the availability of diagnostic tests may further refine the use of early warning capabilities; a more sensitive early detection capability could be combined with a cued use of diagnostic testing to form a much more targeted approach, reducing costs and the likelihood of a false detection. While the results we presented are a first attempt at understanding the potential utility of a host-based early warning system during an outbreak, a comprehensive risk assessment of a QIT policy must consider a range of factors:

- Disease characteristics: prevalence, transmissibility, incubation period, and severity
- Response options: reliable diagnostic tests, vaccines, or treatments
- Resources: cost and availability of QIT measures
- Early warning capability: performance characteristics of early warning systems
- Public health infrastructure: ability to implement an effective public health campaign

An element that we do not include in our model is patient care measures, which are nearly always more effective when deployed earlier than overt indications of infection (e.g., fever). For instance, antiviral drugs (such as zanamivir and oseltamivir/Tamiflu) are most effective in the first ∼48 hours of symptoms [8, 17, 18]; PRESAGED-enabled early warning would allow much faster prescription, use, and potentially more profound therapeutic impact for current dosage recommendations. Triggering the use of diagnostics early would allow clinicians to target drug-based interventions, such as using the precise antibiotic for bacterial infections rather than relying on broad-spectrum versions that contribute to the evolution of drug-resistant bacteria (“superbugs”). Finally, simple supportive care for mild infections, e.g., having rehydrating solutions or over-the-counter symptom-relieving medications available, could ease the burden on health care workers in hospitals, nursing homes, or college campuses. All of these exciting possibilities could be enabled or improved with earlier detection.

Future modeling efforts need to focus on improvements to this model, especially as more is known about the mechanistic basis for host-based early warning of pathogen exposure. Several enhancements of QIT policy include:

1. Consequences of infection, or a cost function of being in the infectious compartment. Adding a more explicit fatality compartment is straightforward. However, more complex models incorporating significantly time-delayed or nonlinear costs are much more accurate for measuring the effectiveness of QIT policies.
2. Early quarantine release. When the maximum incubation period may result in long quarantine periods that lead to unacceptably high quarantine costs, the absence of an exposure detection in individuals who have not been exposed to the pathogen may trigger an early release from quarantine. This early quarantine release approach could significantly reduce costs and civil rights issues associated with quarantine.
3. Modeling of additional QIT responses. Incorporating diagnostic tests into the models could refine the quarantine trigger and release policies. Additionally, future models could include specific treatments upon early warning to modulate infection outcomes. Such options may be particularly important in circumstances when quarantine and isolation resources are limited.
4. Movement among populations. The policy-dependent SEIR model addresses a single, isolated population being homogenously mixed. However, in many circumstances, QIT policy is driven by a concern for the spread of the pathogen into connected populations. To address these circumstances, the policy-dependent SEIR model may be further extended to incorporate changes in each population compartment that result from the movement of people into (or out of) individual population compartments from (or to) other populations. In this formulation, individual populations are modeled with the extended SEIR mode as nodes in a migration network. Network edges are characterized by migration rates between the connected subpopulations.
5. Stochastic modeling. In a more realistic model, each of the SEIR parameters may be modeled in greater detail as a probability distribution rather than a fixed value. Stochastic modeling would support a more comprehensive risk analysis that includes insights about the likely range of potential outcomes, as well as rare but high consequence “edge” cases.

In conclusion, we have shown the epidemiological value of host-based early warning systems in a variety of pathogen outbreaks. By adjusting the underlying assumption, both of the outbreak and the system performance metrics of a notional early detection system, we show in which scenarios early detection is most impactful. The results of this work emphasize the value of early detection in modulating public health responses, though future efforts will also include the value to individual patients.

## Data Availability

None

## Declaration of Interests

The authors declare that they have no competing interests.

## Role of the Funding Source

This effort was funded by Defense Threat Reduction Agency.

The funding sources had no role in writing of the manuscript. The corresponding author had full access to all of the data in the study and had responsibility for submission for publication.

## Notes

### Competing Interest Statement

The authors have declared no competing interest.

### Clinical Trial

No clinical trial conducted

### Funding Statement

This work is funded by the Defense Threat Reduction Agency (DTRA)

